# Hypoxic-Ischemic Encephalopathy Based on clinical biomarkers and associated risk factors in Neonates from Southern Ethiopian Public Hospitals: A Case Control Study

**DOI:** 10.1101/2023.05.28.23290654

**Authors:** Getnet Melaku Ayele, Getachew Mergia, Senait Belay Getahun, Selamawit Semagn Kebede, Zemedu Awoke Ferede, Robel Hussien Kabthymer, Mesfin Abebe

## Abstract

Hypoxic ischemic encephalopathy (HIE) is a serious condition that results from reduced oxygen supply and blood flow to the brain, leading to brain injury and potential long-term neurodevelopmental impairments. This study aimed to identify the maternal and neonatal factors that are associated with HIE among newborns in Ethiopia. An unmatched-control study was conducted in fifteen public hospitals in Southern Nation Nationalities and the Peoples Regional State of Ethiopia. Data were collected from 515 newborns with their index mothers (175 cases and 340 controls) using a structured questionnaire and clinical records which were created and managed by Kobo software for mobile-assisted data collection. Clinical biomarkers were used to diagnose Hypoxic ischemic encephalopathy. Logistic regression analysis was performed to identify the factors associated with Hypoxic ischemic encephalopathy. Maternal education, ultrasound checkup status, gestational age at delivery, mode of delivery, and labor duration were significantly associated with Hypoxic ischemic encephalopathy. Newborns born to illiterate mothers (AOR= 1.913, 95%CI: 1.177, 3.109), those whose mothers did not have an ultrasound checkup during pregnancy (AOR= 1.859, 95%CI: 1.073, 3.221), those who were born preterm (AOR= 4.467, 95%CI: 1.993, 10.012) or post-term (AOR= 2.903, 95%CI: 1.325, 2.903), those who were delivered by cesarean section (AOR= 7.569, 95%CI: 4.169, 13.741), and those who were delivered after prolonged labor (AOR= 3.591, 95%CI: 2.067, 6.238) had higher odds of developing Hypoxic ischemic encephalopathy than their counterparts. This study provides valuable insights into the risk factors for Hypoxic ischemic encephalopathy among newborns in Southern Ethiopia. Improving maternal education and health care services during pregnancy and delivery may help reduce the incidence and severity of Hypoxic ischemic encephalopathy. Future research should use laboratory or imaging investigations, including private health institutions, and explore the mechanisms and outcomes of Hypoxic ischemic encephalopathy.

## Introduction

Hypoxic-ischemic encephalopathy (HIE) is a type of brain dysfunction that occurs when the brain does not receive enough oxygen or blood flow for a period of time [1]. It can cause severe and permanent neurological impairments, such as cerebral palsy, epilepsy, developmental delay, and cognitive impairment which is a leading cause of death or severe disability among infants worldwide[2, 3].

The incidence and risk factors of HIE may vary depending on the geographic region and the level of health care available. In high-income countries, the most common cause of HIE is perinatal asphyxia due to complications during labor and delivery, such as umbilical cord problems, placental abruption, or fetal distress whereas in low- and middle-income countries, it may also result from maternal or fetal infections, anemia, malnutrition, or lack of antenatal care [4, 5].

The global incidence of HIE is 1.5 per 1000 live births, but it varies by region and income level. Low- and middle-income countries have higher rates of HIE (1.8 per 1000 live births) than high-income countries (0.5 per 1000 live births). Within low- and middle-income countries, the incidence of HIE ranges from 1.6 to 7.9 per 1000 live births, with sub-Saharan Africa having the highest rate of 3.1 per 1000 live births and accounting for 30% of the global burden of HIE. In high-income countries, the incidence of HIE ranges from 0.3 to 2.6 per 1000 live births [4, 6].

There is a lack of reliable data on the incidence of HIE in Ethiopia, but some studies have estimated it to be between 10.7 and 30.9 per 1000 live births. The main causes of HIE in Ethiopia are infections in the mother or the baby, long labor, lack of oxygen at birth, and blood infection in the baby. The incidence of HIE in southern Ethiopia is also unclear, but one study found it to be 11.5 per 1000 live births at a tertiary hospital in northern Tanzania, which may be similar to southern Ethiopia [6, 7].

To detect hypoxic-ischemic insult, we can use clinical signs and physical exams as biomarkers. Clinical signs include low Apgar scores, low cord pH, neonatal seizures, and encephalopathy. Physical exams include monitoring brain function, such as level of consciousness, muscle tone, posture, reflexes, and autonomic function. A 5th min APGAR score of <7 or a 10th min APGAR score of <4 also indicate hypoxic-ischemic insult [8-10].

Ethiopia is a low-income country in Sub-Saharan Africa with a high neonatal mortality rate of 23 deaths per 1000 live births in which the prevalence and causes of HIE are not well documented, but some studies have suggested that maternal infections, prolonged labor, birth asphyxia, preterm birth, cord prolapse, post term pregnancy, vacuum mode of delivery, male sex newborn, and neonatal sepsis are common risk factors. In addition, there is also a lack of standardized protocols and resources for the diagnosis and management of HIE in Ethiopian public hospitals [1, 11-14].

Several studies conducted in both developed and developing countries revealed that HIE is predicted by factors related to antepartum, intrapartum, neonatal, and nutritional conditions. Most of the HIE (70.3%) is due to intrapartum factors. Some of the antenatal factors are being a first-time mother, having a previous fetal death/stillbirth, using antidepressants or illicit drugs, having Rh sensitization, and gaining more than 13.6 kg of weight during pregnancy. Some of the intrapartum factors are having a placental abruption, a ruptured uterus, a moderate-to-heavy amount of meconium in the amniotic fluid and having a cesarean-section delivery. These factors are present in 70.3% of the HIE cases [15-19].

Hypoxic-ischemic encephalopathy (HIE) is a serious condition that affects newborns who experience oxygen deprivation and reduced blood flow to the brain which can cause brain damage, seizures, developmental delays, and cerebral palsy [20, 21]. However, the risk factors and biomarkers associated with HIE in neonates from southern

Ethiopian public hospitals are not well understood. Therefore, this study aims to identify the risk factors based on clinical biomarkers associated with HIE in neonates born at public hospitals in Southern Ethiopia. This will help to improve the early detection and treatment of HIE and to reduce the morbidity and mortality associated with this condition.

### Research question

What are the risk factors associated with HIE in neonates from southern Ethiopian public hospitals?

### Hypothesis

Neonates with HIE have higher rates of antenatal, intra-partum, neonatal, and nutritional risk factors than neonates without HIE.

## Materials and Methods

### Study area, study aim and design

An institution-based unmatched case-control study was conducted in public hospitals of SNNPR, Ethiopia from June 2022 to March 2023 to identify risk factors based on clinical biomarkers associated with Hypoxic-Ischemic Encephalopathy in Neonates.

### Study population and Methods

The study population consisted of neonates born at public hospitals in SNNPR. Cases were neonates who had HIE diagnosed at public hospitals in Southern Ethiopia during the study period, while controls were neonates who did not have HIE diagnosed in the same area and period. Newborns that had any condition that made them unable to survive outside the womb were excluded from the study.

We considered the following exposure variables to calculate the sample size: maternal anemia, fetal presentation, color of amniotic fluid and mode of delivery. We chose maternal anemia as an independent variable because it yielded the largest sample size compared to other exposure variables. We used Open Epi version 7 statistical software package and a formula for two population proportions to calculate the sample size. Based on a previous study in southern Ethiopia, we assumed that 6.74% of the controls (newborns who had adequate breathing and/or cry and APGAR score >7) were exposed [22].

We made the following assumptions for the sample size calculation: 17.98% of the cases were exposed, a 95% CI, 80% power of the study, and a 2:1 ratio of controls to cases with a 3.87 odds ratio. This resulted in a sample size of 243 newborns (81 cases and 162 controls). However, since the study was regional and had multiple stages, we applied a design effect and doubled the sample size to 486 newborns (162 cases and 324 controls). We also accounted for a 10% nonresponse rate and aimed to study **535** newborns (178 cases and 357 controls).

We selected 15 out of 79 government hospitals in SNNPR by proportionally allocating samples to each hospital based on the average case flow estimated by enrollment. We used well-designed data collection tools and trained data collectors on how to observe deliveries, APGAR scoring, clinical signs of HIE and fill data to ensure data quality. We created the instrument by reviewing various sources of literature. To ensure its accuracy and clarity, we translated it from English to Amharic and then back to English. We used a pre-tested structured questionnaire to collect data from the participants [7, 23, 24].

We used Kobo toolbox, a free and open-source application, to design, manage and collect data for our questionnaire. Kobo toolbox allows us to collect data online or offline using mobile devices, and to analyze and visualize data easily. We selected Kobo toolbox because of its user-friendly, secure, and reliable interface. It also offers features that enhance data quality control, such as validation rules and skip logic. Our data collectors were professional midwives and nurses who worked at different hospitals from where they collected data.

### Operational definitions

#### Hypoxic-ischemic encephalopathy (HIE)

A type of brain dysfunction that occurs when a newborn does not receive enough oxygen and blood flow during or after birth.

#### Diagnosis of HIE

To diagnose HIE, the researcher used clinical signs and physical exams. The clinical signs were low Apgar scores (below 7 at 5 minutes or below 4 at 10 minutes) and neonatal seizures. The physical exams evaluated consciousness, muscle tone, posture, reflexes, and autonomic function. **Clinical Biomarkers:** The researcher used the following clinical biomarkers to indicate HIE: Apgar scores, neonatal seizures, level of consciousness, muscle tone, posture, reflexes, and autonomic function. **Cases:** Newborns who were delivered and diagnosed with Hypoxic-ischemic encephalopathy were considered as cases. **Controls:** Newborns who were delivered and did not have hypoxic-ischemic encephalopathy (normal).

### Data processing and analysis

We used SPSS version-26 to enter and analyze the data. We ensured the data quality and performed descriptive and inferential statistics. We used cross-tabulation and logistic regression to assess the relationship between dependent and independent variables. We controlled confounding factors by using adjusted odds ratio (AOR). We set the significance level at 0.05 and the confidence level at 95%.

### Ethics approval and consent to participate

This research followed the Declaration of Helsinki’s recommendation to consider the potential benefits for the participants [25, 26]. Studying the risk factors of HIE involves ethical and methodological issues that are specific to human subjects research. Therefore, this study adhered to the WHO’s ethical and safety guidelines for human subjects’ research [27]. The Institutional Review Board (IRB) of Dilla University, College of Medicine and Health

Sciences approved this study ethically (protocol unique number 010/18-06) and wrote an official support letter to the regional health department of SNNPR. The regional health department also gave an official letter to each selected hospital. Moreover, the index mothers of the neonates who participated in the study gave their written consent after being informed about the study. The study participants were free to withdraw from the study at any point during data collection.

## Results

### Maternal Socio-demographic and reproductive characteristic

This study included 175 neonates with Hypoxic Ischemic Encephalopathy (HIE) (cases) and their index mothers, and 340 neonates without Hypoxic Ischemic Encephalopathy (controls) and their index mothers, with a response rate of 96.26%. The mean age of the mothers in the study was 26.57 years (+SD=4.317). The occupational status of the index mother showed that more than forty-eight percent of the mothers who delivered babies with HIE (cases) and more than thirty percent of the mothers whose newborns were normal (controls) were unemployed. Regarding the duration of the current pregnancy, most of the mothers (54.9% of cases and 52.1% of controls) delivered their babies at term (38-42 weeks of gestation). However, a significant proportion of mothers (18.9% of cases and 17.1% of controls) experienced some illness during their current pregnancy **(Table 1)**.

**Table 1:** Maternal Socio-Demographic and Reproductive Characteristics of Newborns Delivered at Public Hospitals in Southern Ethiopia, 2023 (n=515)

| Variable | Category | Case n=175 (%) | Control n=340 (%) | Total n=515 (%) |
| --- | --- | --- | --- | --- |
| Occupation of index mother | Unemployed | 85 (48.6) | 104 (30.6) | 189 (36.7) |
|  | Employed | 90 (51.4) | 236 (69.4) | 326 (63.3) |
| Maternal educational status | Literate | 99 (56.6) | 252 (74.1) | 351 (68.2) |
|  | Illiterate | 76 (43.4) | 88 (25.9) | 164 (31.8) |
| Pregnancy status | Singleton | 94 (53.7) | 196 (57.6) | 290 (56.3) |
|  | Multiple (>2) | 81 (46.3) | 144 (42.4) | 225 (43.7) |
| Parity | Primi | 68 (38.9) | 109 (32.1) | 177 (34.4) |
|  | Multi | 107 (61.1) | 231 (67.9) | 338 (65.6) |
| Duration of current pregnancy | <= 37 weeks | 66 (37.7) | 105 (30.9) | 171 (33.2) |
|  | 38 - 42 weeks | 96 (54.9) | 177 (52.1) | 273 (53.0) |
|  | 42+ weeks | 13 (7.4) | 58 (17.1) | 71 (13.8) |
| History of bad pregnancy outcome | Miscarriage | 58 (33.1) | 129 (37.9) | 187 (36.3) |
|  | Still birth | 87 (49.7) | 119 (35.0) | 206 (40.0) |
|  | Child death | 30 (17.1) | 92 (27.1) | 122 (23.7) |

### Prenatal Factors Associated with Hypoxic Ischemic Encephalopathy

This study examined antepartum, intra-partum, nutritional and neonatal factors. During their current pregnancy, about nineteen percent of cases and eighteen percent of controls had some illness among the index mothers who participated in the study. Of these, 36.4% of cases and 32.2% of controls were anemic during their antepartum period. Concerning the ANC visit status of the index mothers, 82 (59.9%) cases and 154 (51.2%) controls did not finish their fourth ANC visit. Moreover, 68 (89.5%) of mothers whose babies had HIE (cases) and 191 (80.9%) of mothers whose babies were normal (without HIE diagnosis or controls) had normal amniotic fluid volume **(Table 2)**.

**Table 2:** Prenatal Factors Associated with Hypoxic Ischemic Encephalopathy among Newborns Delivered at Public Hospitals in Southern Ethiopia, 2023 (n=515)

| Variable | Category | Case n=175 (%) | Control n=340 (%) | Total n=515 (%) |
| --- | --- | --- | --- | --- |
| Illness during pregnancy? | Yes | 33 (18.9) | 58 (17.1) | 91 (17.1) |
|  | No | 142 (81.1) | 282 (82.9) | 424 (82.3) |
| Types of Illness during pregnancy | Hypertension | 13 (39.4) | 34 (57.6) | 47 (51.1) |
|  | Anemia | 12 (36.4) | 19 (32.2) | 31 (33.7) |
|  | Bleeding | 8 (24.2) | 6 (10.2) | 14 (15.2) |
| ANC visits during pregnancy | Yes | 137 (78.3) | 301 (88.5) | 438 (85.0) |
|  | No | 38 (21.7) | 39 (11.5) | 77 (15.0) |
| ANC visit status | Completed ANC 4th | 55 (40.1) | 147 (48.8) | 202 (46.1) |
|  | Not Completed ANC 4th | 82 (59.9) | 154 (51.2) | 236 (53.9) |
| Ultrasound check up during your time of pregnancy | Yes | 91 (52.0) | 234 (68.8) | 325 (63.1) |
|  | No | 84 (48.0) | 106 (31.2) | 190 (36.9) |
| Amniotic fluid status checkup during pregnancy time | Yes | 76 (43.4) | 236 (69.4) | 312 (60.6) |
|  | No | 99 (56.6) | 104 (30.6) | 203 (39.4) |
| The status of amniotic fluid | Polyhydramnios | 4 (5.3) | 25 (10.6) | 29 (9.3) |
|  | Oligohydramnios | 4 (5.3) | 20 (8.5) | 24 (7.7) |
|  | Normal | 68 (89.5) | 191 (80.9) | 259 (83.0) |

### Intra-partum Factors Associated with Hypoxic Ischemic Encephalopathy (HIE)

We assessed many intrapartum-related variables at the time of labor and delivery. We found that most of the neonates with HIE (cases) (86.9%) and without HIE (controls) (98.2%) had a cephalic presentation. About 40.0% of cases and 8.2% of controls were delivered by cesarean section (C/S) mode of delivery and among those delivered vaginally, 62.9% of cases and 92.6% of controls had spontaneous labor. We also checked if the mothers who were in labor took any drugs. We found that 33.1% of cases and 26.4% of controls took some anti-pain or anesthetic drug. Regarding the duration of labor and rupture of the membrane during the intrapartum period, more than half of the cases (53.1%) and more than a third of the cases (34.9%) had more than 12 hours of duration, respectively. Of all the study participants, 41.1% of cases and 14.4% of controls encountered labor and delivery-related complications. The status of amniotic fluid was also an important variable, with 50.3% of cases and 85.3% of controls having clear amniotic fluid status, which represented 73.4% of all index mothers **(Table 3)**.

**Table 3:**
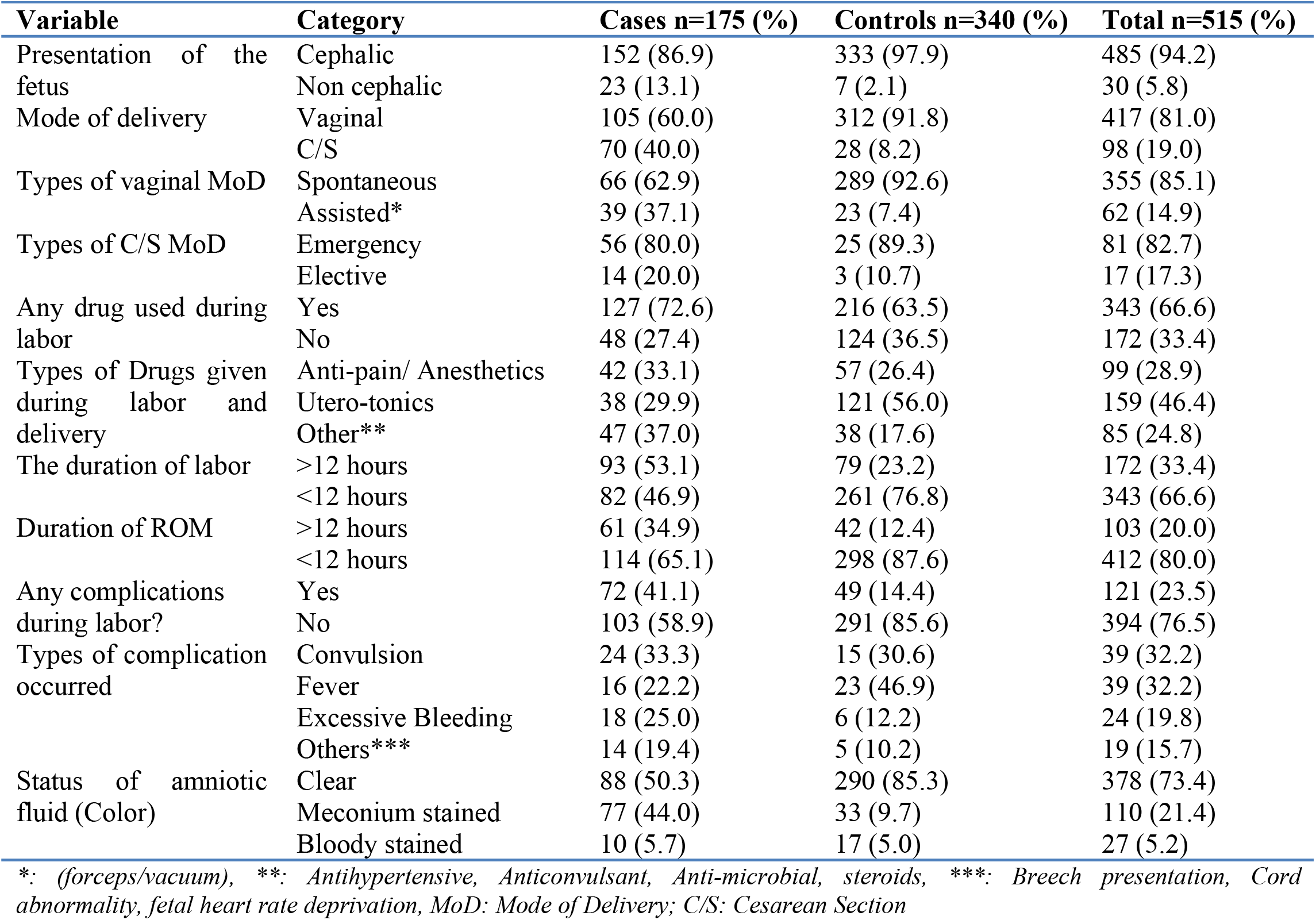
Intra-partum Factors Associated with Hypoxic Ischemic Encephalopathy among Newborns Delivered at Public Hospitals in Southern Ethiopia, 2023 (n=515)

| Variable | Category | Cases n=175 (%) | Controls n=340 (%) | Total n=515 (%) |
| --- | --- | --- | --- | --- |
| Presentation of the fetus | Cephalic | 152 (86.9) | 333 (97.9) | 485 (94.2) |
|  | Non cephalic | 23 (13.1) | 7 (2.1) | 30 (5.8) |
| Mode of delivery | Vaginal | 105 (60.0) | 312 (91.8) | 417 (81.0) |
|  | C/S | 70 (40.0) | 28 (8.2) | 98 (19.0) |
| Types of vaginal MoD | Spontaneous | 66 (62.9) | 289 (92.6) | 355 (85.1) |
|  | Assisted* | 39 (37.1) | 23 (7.4) | 62 (14.9) |
| Types of C/S MoD | Emergency | 56 (80.0) | 25 (89.3) | 81 (82.7) |
|  | Elective | 14 (20.0) | 3 (10.7) | 17 (17.3) |
| Any drug used during labor | Yes | 127 (72.6) | 216 (63.5) | 343 (66.6) |
|  | No | 48 (27.4) | 124 (36.5) | 172 (33.4) |
| Types of Drugs given during labor and delivery | Anti-pain/ Anesthetics | 42 (33.1) | 57 (26.4) | 99 (28.9) |
|  | Utero-tonics | 38 (29.9) | 121 (56.0) | 159 (46.4) |
|  | Other** | 47 (37.0) | 38 (17.6) | 85 (24.8) |
| The duration of labor | >12 hours | 93 (53.1) | 79 (23.2) | 172 (33.4) |
|  | <12 hours | 82 (46.9) | 261 (76.8) | 343 (66.6) |
| Duration of ROM | >12 hours | 61 (34.9) | 42 (12.4) | 103 (20.0) |
|  | <12 hours | 114 (65.1) | 298 (87.6) | 412 (80.0) |
| Any complications during labor? | Yes | 72 (41.1) | 49 (14.4) | 121 (23.5) |
|  | No | 103 (58.9) | 291 (85.6) | 394 (76.5) |
| Types of complication occurred | Convulsion | 24 (33.3) | 15 (30.6) | 39 (32.2) |
|  | Fever | 16 (22.2) | 23 (46.9) | 39 (32.2) |
|  | Excessive Bleeding | 18 (25.0) | 6 (12.2) | 24 (19.8) |
|  | Others*** | 14 (19.4) | 5 (10.2) | 19 (15.7) |
| Status of amniotic fluid (Color) | Clear | 88 (50.3) | 290 (85.3) | 378 (73.4) |
|  | Meconium stained | 77 (44.0) | 33 (9.7) | 110 (21.4) |
|  | Bloody stained | 10 (5.7) | 17 (5.0) | 27 (5.2) |

\*. (forceps/vacuum), \*\*: Antihypertensive, Anticonvulsant, Anti-microbial, steroids, \*\*\*: Breech presentation, Cord abnormality, fetal heart rate deprivation, MoD: Mode of Delivery; C/S: Cesarean Section

### Nutritional related factors of neonatal hypoxic ischemic encephalopathy

This study assessed the nutritional status of the mothers of newborns. The following figure clearly demonstrates that meat and meat products were the most preferred food among the pregnant mothers of the newborns in our study. In fact, more than half of the cases (58.3%) and the controls (50.0%) chose this food category over any other option **(Fig 1)**.

**Figure 1:**
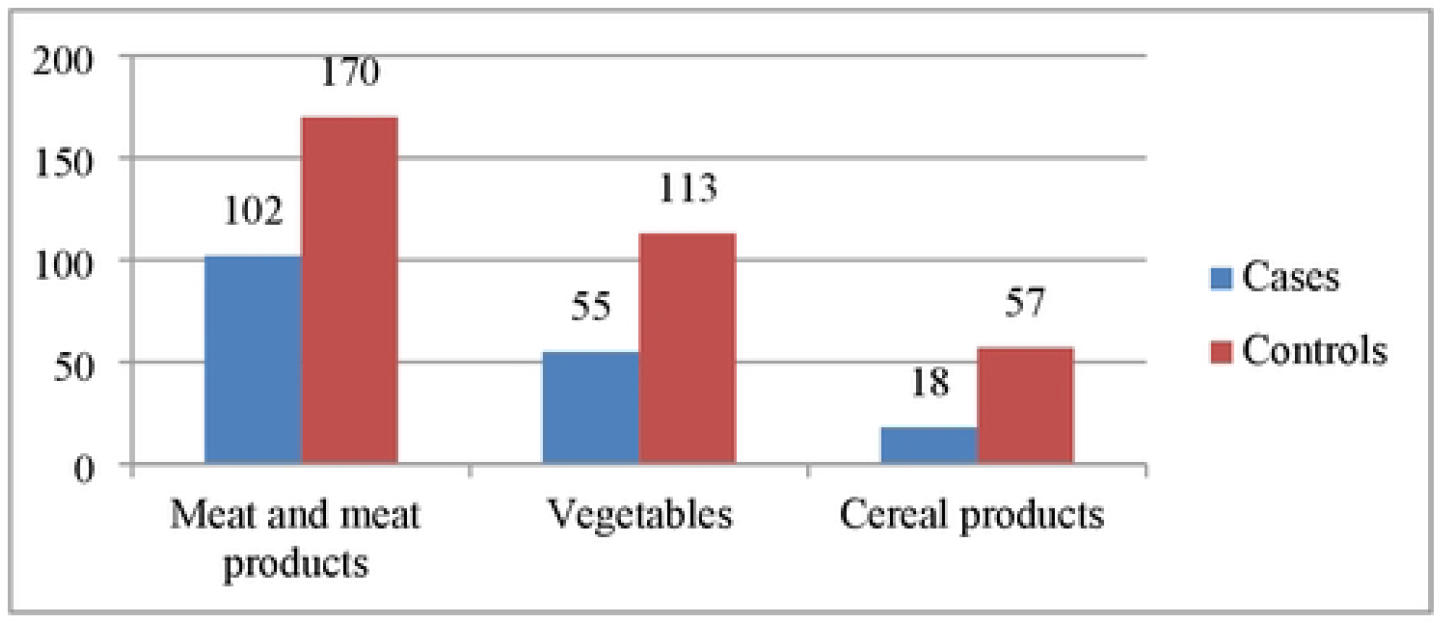
Preferred Foods of Pregnant Mothers of Newborns Delivered at Public Hospitals in Southern Ethiopia, 2023 (n=515)

We found that 96% of the cases and 98% of the controls had a feeding frequency of more than three times per day. However, only 35.4% of the mothers whose newborns had HIE (cases) and 47.9% of the mothers whose newborns did not have HIE (controls) did not have any knowledge about the balanced diet recommended during pregnancy. Moreover, 62.9% of the cases and 80.3% of the controls were unaware that some foods are not recommended during pregnancy.

### Neonatal factors associated with neonatal hypoxic ischemic encephalopathy

The neonates delivered had a balanced gender distribution. The average APGAR score was 7.06 (+SD = 1.444), indicating a good health status. The average birth weight of the newborns was 2873.2 grams (±SD = 480.171 grams), and most of them, 66.3% of the cases and 80.0% of the controls, were within the normal range of 2501 - 3999 grams **(Fig 2)**. We also assessed intrauterine growth restriction (IUGR) immediately after delivery, and found that only a few neonates, 5.1% of the cases and 4.7% of the controls, had IUGR.

**Figure 2:**
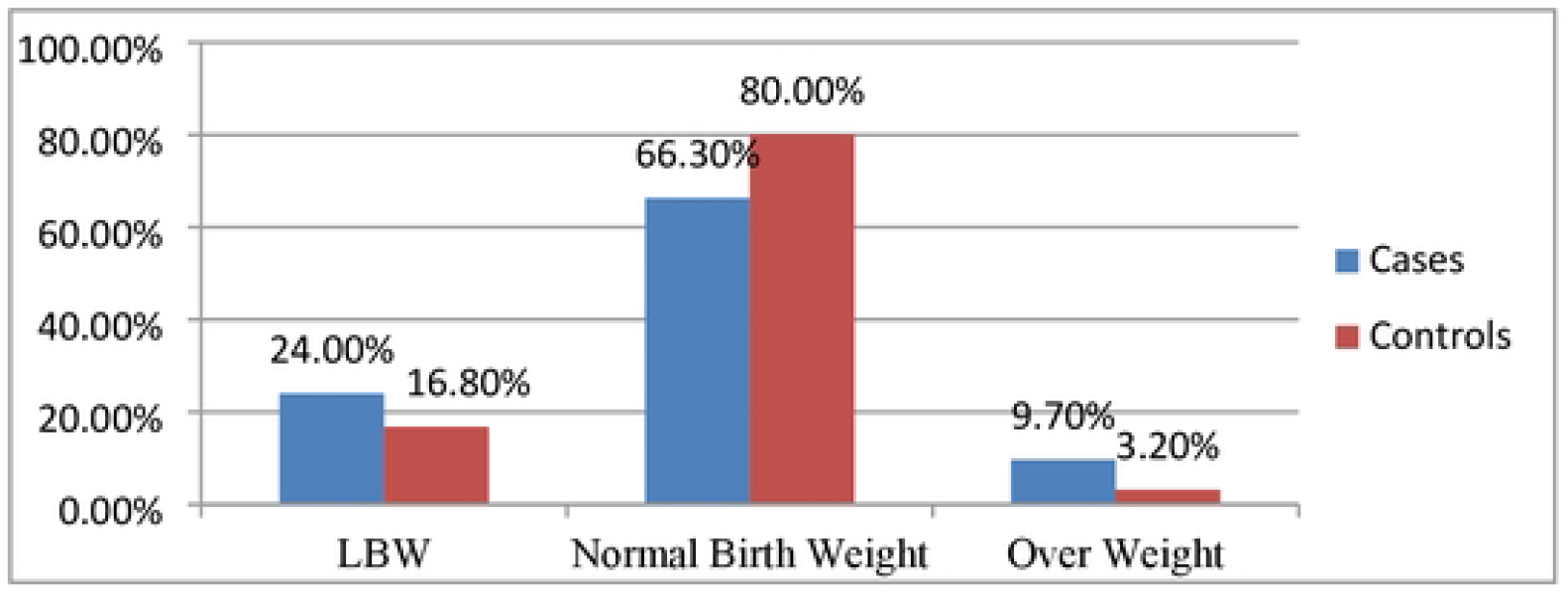
Birth weight of the newborns Delivered at Public Hospitals in Southern Ethiopia, 2023 (n=515)

### Factors associated with the occurrence of neonatal Hypoxic Ischemic Encephalopathy

To adjust for potential confounders, we performed a multivariable analysis with the following predictor variables that had a p-value of less than 0.2 in the binary logistic regression: index mother’s occupation and education level, gestational age at delivery, ANC attendance during pregnancy, ultrasound examination during pregnancy, mode of delivery, drug use during labor, labor duration, ROM duration, index mother’s knowledge of recommended foods during pregnancy and newborn’s birth weight.

The multivariable logistic regression analysis model showed that the following variables were statistically significant: index mother’s education level, gestational age at delivery, ultrasound checkup status during pregnancy, mode of delivery and labor duration. Neonates born to illiterate index mothers had more than twice the odds (AOR= 1.913, 95%CI: 1.177, 3.109) of developing hypoxic ischemic encephalopathy than those born to literate mothers.

We assessed gestational age at delivery and found that preterm newborns (born before 37 weeks of gestation) had nearly 4.5 times the odds (AOR= 4.467, 95%CI: 1.993, 10.012) and post-term newborns (born after 42 weeks of gestation) had nearly 3 times the odds (AOR= 2.903, 95%CI: 1.325, 2.903) of developing hypoxic-ischemic encephalopathy compared with term newborns (born between 38-42 weeks of gestation).

We also examined the effect of ultrasound examination during pregnancy and delivery method on hypoxic-ischemic encephalopathy. We found that newborns whose mothers did not have an ultrasound examination during pregnancy had twice the odds (AOR= 1.859, 95%CI: 1.073, 3.221) of developing hypoxic-ischemic encephalopathy than those whose mothers did. Moreover, newborns delivered by cesarean section (C/S) had more than seven times the odds (AOR= 7.569, 95%CI: 4.169, 13.741) of suffering from hypoxic-ischemic encephalopathy than those delivered vaginally. We also found that labor duration was associated with hypoxic-ischemic encephalopathy. Newborns delivered after prolonged labor (≥12 hours) had 3.6 times the odds (AOR= 3.591, 95%CI: 2.067, 6.238) of developing hypoxic-ischemic encephalopathy than those delivered after normal labor duration (≤12 hours) ***(Table4)***.

**Table 4:**
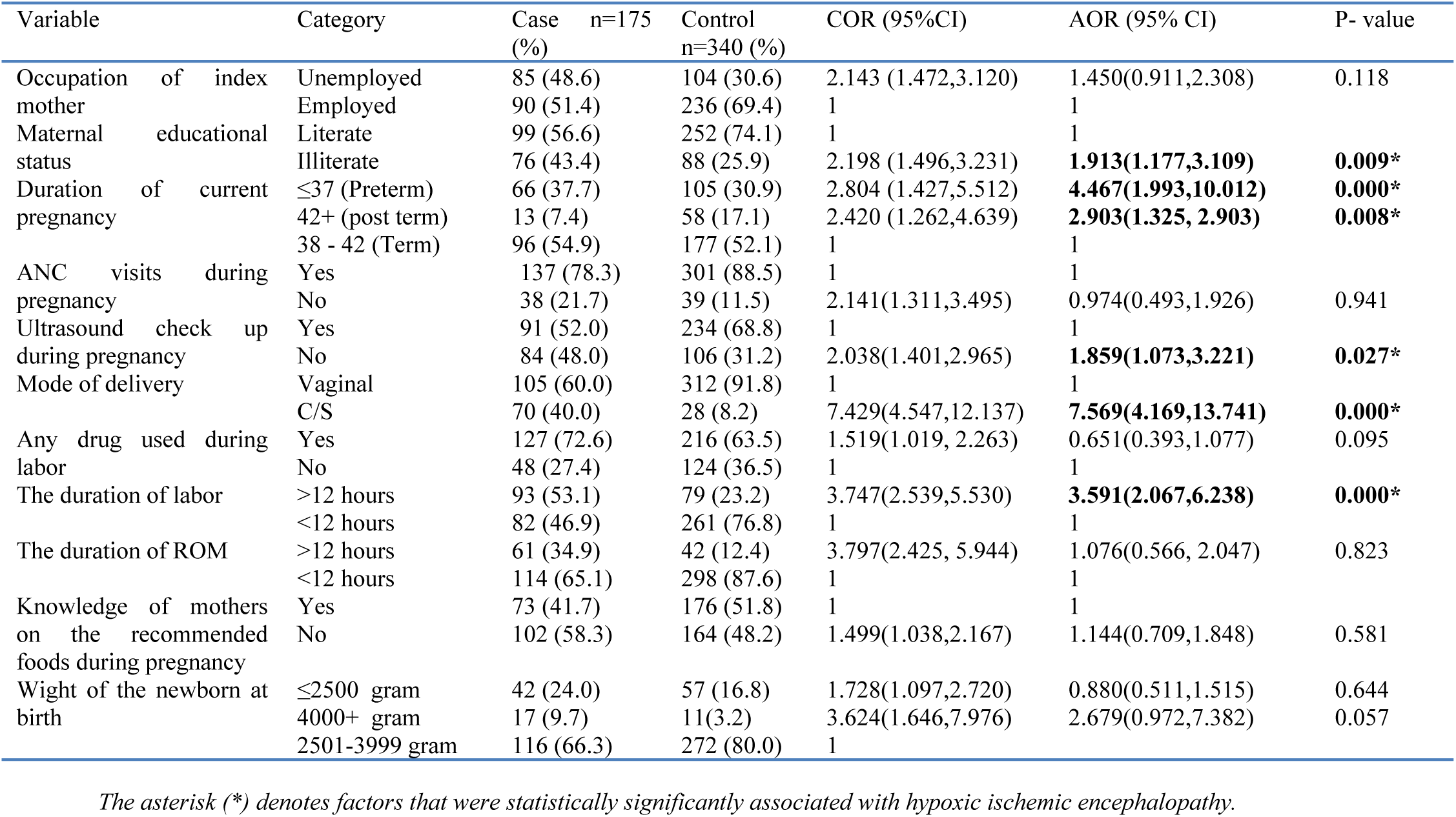
Bivariate and Multivariable Logistic Regression among Newborns Delivered at Public Hospitals in Southern Ethiopia, 2023 (n=515)

## Discussion

This study identified several maternal and neonatal factors that were associated with increased risk of hypoxic ischemic encephalopathy (HIE) among newborns which is a serious condition that results from reduced oxygen supply and blood flow to the brain, leading to brain injury and potential long-term neurodevelopmental impairments [28]. The findings of this study have important implications for the prevention and management of HIE.

One maternal factor significantly associated with HIE was the index mother’s education level. Newborns born to illiterate mothers had two times the odds of developing HIE than those born to literate mothers. This result is in line with a study conducted in Northern and Southern Ethiopia [7, 29]. The finding suggests that maternal literacy may influence the awareness and utilization of antenatal and intra-partum care services, which are essential for detecting and preventing complications that may cause HIE [30]. Maternal literacy may also affect the knowledge and practice of healthy behaviors during pregnancy, such as nutrition, hygiene, and infection prevention [18, 31].Therefore, improving maternal education and empowerment may be a key strategy to reduce the risk of HIE among newborns.

This study identified ultrasound checkup status during pregnancy and gestational age at delivery as maternal factors associated with HIE. Newborns whose mothers did not have an ultrasound checkup during pregnancy had higher odds of developing HIE than those whose mothers did, which is in line with previous studies done in Cameroon and Istanbul [18, 32]. Ultrasound checkup is a valuable tool for detecting and preventing conditions that may cause HIE, such as fetal anomalies, growth restriction, placental abnormalities, and abnormal fetal presentation. Ultrasound checkups may also help determine the gestational age and the optimal timing and mode of delivery, which are essential for avoiding preterm and post-term births. Both preterm and post-term births were found to increase the risk of HIE in this study, as well as in other studies done in Australia, Northern Ethiopia, and Addis Ababa [7, 29, 33, 34].

Preterm newborns (born before 37 weeks of gestation) had 4.5 times the odds of developing HIE, while post-term newborns (born after 42 weeks of gestation) had 3 times the odds of developing HIE compared with term newborns. These findings are consistent with recent studies that have suggested that post-term birth may be associated with placental insufficiency, fetal distress, meconium aspiration, and birth trauma, which may compromise the oxygen supply and blood flow to the brain [35, 36]. Therefore, this study recommends ensuring access and availability of ultrasound checkups during pregnancy and monitoring gestational age at delivery as effective measures to prevent HIE.

The mode of delivery was another factor that influenced the risk of HIE. Newborns delivered by cesarean section (C/S) had more than 7 times the odds of developing HIE than those delivered vaginally. This is consistent with several studies conducted in Cameroon, Istanbul, and Northern Ethiopia [18, 19, 37]. This finding may reflect the fact that C/S is often performed as an emergency intervention when there is fetal distress or other complications that may compromise the oxygen supply to the fetus. However, C/S may also pose some risks for the newborn, such as respiratory distress syndrome, transient tachypnea, and iatrogenic prematurity [38]. Therefore, C/S should be reserved for cases where there is a clear indication and benefit for the mother and the fetus and performed with adequate monitoring and care to minimize the risk of HIE.

The labor duration was another factor that affected the risk of HIE. Newborns delivered after prolonged labor (≥12 hours) had 3.6 times the odds of developing HIE than those delivered after normal labor duration (≤12 hours). This finding is consistent with previous studies conducted in developed and developing countries that have shown that prolonged labor is associated with increased risk of fetal hypoxia, acidosis, meconium aspiration, infection, and birth trauma [7, 39, 40]. Prolonged labor may also result from cephalopelvic disproportion or malposition of the fetus, which may require instrumental delivery or C/S [41]. Therefore, timely detection and management of prolonged labor is essential for preventing HIE and its sequelae.

This study has some limitations that warrant further discussion. First, it relied on clinical biomarkers without laboratory or imaging investigation, which could introduce some subjectivity bias in the diagnosis. Second, it excluded private health institutions, health centers and health posts that offer substantial maternal and child health related services due to budget constraints, which could limit its applicability to other populations or contexts. Third, it did not evaluate the severity or outcome of HIE among newborns, which could vary depending on the extent and location of brain damage.

Despite these limitations, this study provides valuable insights into the risk factors for HIE among newborns in Ethiopia. It was regional and included most tertiary care hospitals that have a wide catchment population and used a relatively large sample size. The findings suggest that improving maternal education and health care services during pregnancy and delivery may help reduce the incidence and severity of HIE. Future research should explore the mechanisms and pathways by which these factors influence the risk of HIE, as well as the long-term outcomes and interventions for newborns with HIE.

## Conclusion

This study aimed to identify the maternal and neonatal factors that are associated with hypoxic ischemic encephalopathy (HIE) among newborns in Southern Ethiopia. HIE is a serious condition that results from reduced oxygen supply and blood flow to the brain, leading to brain injury and potential long-term neurodevelopmental impairments. The study found that maternal education, ultrasound checkup status, gestational age at delivery, mode of delivery, and labor duration were significantly associated with HIE. Newborns born to illiterate mothers, those whose mothers did not have an ultrasound checkup during pregnancy, those who were born preterm or post-term, those who were delivered by cesarean section, and those who were delivered after prolonged labor had higher odds of developing HIE than their counterparts. These findings suggest that improving maternal education and health care services during pregnancy and delivery may help reduce the incidence and severity of HIE. This study recommends future research to use laboratory or imaging investigations, including private health institutions, and explore the mechanisms and outcomes of HIE. It also recommends health care providers to ensure ultrasound checkup, monitor gestational age and mode of delivery, and detect and manage prolonged labor.

## Data Availability

Availability of Data and Materials: The data that supports this study is not publicly available due to ethical and privacy concerns. However, the corresponding author will share the data upon reasonable request. The data include anonymized demographic and clinical data of the participants and questionnaires. The data will be accessible for 10 years after the publication of this paper. To request data access, please contact Getnet Melaku Ayele at.

## Abbreviations and acronyms

ANC: Antenatal Clinic
AOR: Adjusted Odds Ratio
APGAR: Activity-Pulse-Grimace-Appearance-Respiration
C/S: Cesarean Section
CI: Confidence Interval
HIE: Hypoxic Ischemic Encephalopathy
IRB: Institutional Review Board
IUGR: Intrauterine Growth Restriction
LBW: Low Birth Weight
ROM: Rupture of Membrane
SD: Standard Deviation
SNNPR: South Nation Nationalities People’s Region

## Acknowledgements

We are deeply grateful to the SNNPR of Ethiopia health department for their cooperation and permission to conduct this study in the area. We also appreciate the personnel of selected hospitals and the women with their neonate who participated in this study for their invaluable contributions. Moreover, we thank the data collectors and supervisors for their role in data collection and the anonymous reviewers for their constructive comments and suggestions that enhanced the quality of the manuscript.

